# Regional Prevalence of Hypertension Among People Diagnosed with Diabetes in Africa, A Systematic Review and Meta-analysis

**DOI:** 10.1101/2023.04.26.23289171

**Authors:** Thomas Hinneh, Samuel Akyirem, Irene Fosuhemaa Bossman, Victor Lambongang, Patriot Ofori-Aning, Oluwabunmi Ogungbe, Yvonne Commodore Mensah

**Affiliations:** Johns Hopkins University School of Nursing; Yale School of Nursing, Yale University, USA; School of Health and Life Sciences, Glasgow Caledonian University, Scotland; School of Health Sciences, Liberty University, USA; Department of Medicine for Older People, Stockport NHS Foundation Trust; Johns Hopkins Bloomberg School of Public Health

**Keywords:** hypertension, diabetes, Co-morbidity, non-communicable diseases, Africa

## Abstract

**Background:** Hypertension and diabetes share common pathophysiological mechanisms and hence have a high likelihood of co-occurring. The co-existence of hypertension and diabetes increases cardiovascular disease risk and healthcare spending. This review aimed to estimate the burden of hypertension among people with diabetes in African countries.

**Methods:** This review was registered in the International Prospective Register of Systematic Reviews (CRD42021256221). We searched PubMed, Embase, and Hinari databases to identify peer-reviewed articles which provided data on the prevalence of hypertension in people diagnosed with diabetes in Africa. Studies included in the review used different diagnostic criteria and thresholds for hypertension and diabetes diagnosis. We quantified the prevalence of hypertension using random-effects models. We applied generalized linear mixed models with logit transformation to compute regional and overall pooled prevalence and estimate heterogeneity (I^2^).

**Results:** Out of 3810 studies retrieved from various sources, 41 met the inclusion criteria with sample sizes ranging from 80 - 116726. The mean age was 58 (± 11) years and 56% were women. The pooled prevalence of hypertension in people diagnosed with diabetes was 58.1% [95% CI: 52.0% - 63.2%]. By African region, Central Africa had the highest hypertension prevalence; 77.6% [95% CI: 53.0% - 91.4%], South Africa 69.1% [95% CI: 59.8% - 77.1%;], North Africa 63.4% [95% CI: 37.1% - 69.1%;], West Africa 51.5% [95% CI: 41.8% - 61.1%] and East Africa 53.0% [95% CI: 45.8% - 59.1%]. Increasing age, being overweight/obese, being employed, longer duration of diabetes, urban residence, and male sex were associated with a higher likelihood of hypertension diagnosis (p-values<0.005).

**Conclusion:** The high prevalence of hypertension among people with diabetes in Africa, highlights the critical need for an integrated differentiated service delivery to improve and strengthen primary care and prevent cardiovascular disease. Findings from this meta-analysis may inform the delivery of interventions to prevent premature cardiovascular disease deaths among persons in African countries.

## Introduction

Hypertension and diabetes are two of the most prevalent non-communicable diseases (NCDs), and major causes of morbidity and mortality globally (1). The burden of hypertension has risen substantially to over one billion since 1990, according to the Lancet NCD Risk Factor Collaboration Study (NCD-RisC) (1). During the same period (1990-2017), the global prevalence of diabetes also increased from 211.2 million to 476 million (2). Nearly two-thirds of hypertension and diabetes cases occur in low- and middle-income countries (LMICs), where health systems are already burdened with infectious diseases (3). Even though the Lancet NCD-RisC study reported a decline in the burden of hypertension in high-income countries, LMICs continue to experience an upsurge in rates of hypertension (2–4).

Diabetes and hypertension have a higher likelihood of co-occurring because they share several common risk factors including low physical activity, unhealthy dietary patterns, and obesity (5). It is therefore not surprising that people who develop diabetes are also at a greater risk for developing hypertension and vice versa. Up to 60% of patients with hypertension are likely to develop diabetes (6). In a nationally representative survey conducted in Bangladesh, hypertension-diabetes co-morbidity affected 4.7% of the population and the prevalence was significantly higher among educated and unemployed people aged 70 years and above (7). In a Korean study, persons with hypertension had a 51% higher risk of developing type 2 diabetes (5). However, data on the burden of hypertension among people diagnosed in Africa is limited despite the link between diabetes and hypertension.

Hypertension and diabetes are also linked pathophysiologically. Diabetes causes the stiffening of small blood vessels, and arteriosclerosis which elevates peripheral vascular resistance and significantly increases the risk of developing hypertension (8). Similarly, hypertension increases insulin resistance, therefore, increasing patients’ risk of developing diabetes and complications (6).

Co-morbidity of hypertension and diabetes worsens blood pressure control and quality of life (9–11), increases the economic burden on patients, and complicates healthcare needs. Persons living with both hypertension and diabetes are less likely to receive appropriate care until the occurrence of complications (12). Hypertension and diabetes co-morbidity may significantly affect the quality of care as well as glucose and blood pressure control (13). This underscores the need for an equal prioritization of clinical and community-based interventions for hypertension and diabetes given their strong association with the risk of developing cardiovascular diseases.

An Integrated management strategy may provide a platform for cost-effective management of NCDs and associated risk factors in Africa. The concept of integrated management of NCDs framework is part of the WHO Package of essential noncommunicable (PEN) disease strategies to provide a common treatment platform, improve treatment outcomes and enhance equity and efficiency in healthcare delivery. However, there is a need to first understand the magnitude of hypertension and diabetes co-morbidity in Africa by quantifying the prevalence of hypertension among people diagnosed with diabetes to inform the planning and delivery of an appropriate NCD care model in Africa. Therefore, this review seeks to synthesis the burden of hypertension among people diagnosed with diabetes in Africa using meta-analytic methods.

## Methods

This study was conducted according to a pre-designed protocol and the Preferred Reporting Items for a Systematic Review and Meta-analysis (PRISMA) guideline (14). The review was registered on the International Prospective Register of Systematic Reviews (Prospero Registration ID: CRD42021256221)(15).

### Search strategy

A comprehensive literature search was performed on PubMed, Embase, and HINARI databases to identify relevant articles that provided data on the prevalence or incidence of hypertension among people diagnosed with diabetes in African countries. The search terms were categorised into population, outcome, and region. Specific terms developed under each category were (1) population: adults diagnosed with type 1 and type 2 diabetes mellitus, with or without hypertension, (2) Outcome: prevalence, incidence, risk factors, co-morbidity, treatment outcomes of diabetes and/or hypertension, and (3) Region: Africa. Based on these pre-specified search terms, a search string for: “Diabetes type 1” OR “Diabetes type 2” AND “hypertension” OR “high blood pressure” AND “Incidence” OR “prevalence” OR “co-morbidity” OR “risk factors” OR “treatment outcomes” AND “Africa” OR “All African countries” was derived through an iterative process and adapted for all the databases. Where applicable Medical Subject Headings (MeSH) together with Boolean operators (“AND” and “OR”) were applied. This research was not focused on trends, so this term was not included in the search strategy. The original search covered from inception to September 2021 on each database. An updated search was conducted on March 23, 2023, to cover the entire period from inception to March 2023. This review is the first study to estimate the prevalence of hypertension among people with diabetes and so no limitation was applied in terms of year of publication. The full search strategies are available **(Supplementary File 1)**.

### Eligibility criteria

We used the ‘‘Population, outcome, and region’’ strategy to guide the inclusion and exclusion criteria. Population included studies that recruited participants diagnosed with diabetes type 1 or type 2 were included. The main outcome of the review was the reported prevalence or incidence of hypertension among people with diabetes. Studies that reported secondary-related outcomes such as pregnancy-induced hypertension were excluded. Hypertension threshold was specified as ≥140/90 mm Hg or ≥130/80 mm Hg given that cut-off varies across countries in Africa. Studies conducted in African countries were included in the review.

### Study selection and data extraction

All the articles retrieved from the various databases were exported to Endnote Version X 9.0 software and duplicates were removed. Two reviewers, IB and VL independently conducted title and abstract screening using the Rayyan Software platform. TH and AS resolved any disagreements when the two reviewers could not reach a consensus. TH, OP, and AS independently extracted the data from the included articles using Microsoft Office Excel. Data items including study title, authors, date of publication, region, country, type of participants, age, and the reported outcome were extracted. Other outcomes date related to hypertension such as prevalence or incidence of hypertension, complications, diagnostic thresholds, and associated risk factors, were also extracted.

### Synthesis of results

We performed a tabular synthesis of sample characteristics, effect measures (prevalence), main findings, diagnostic threshold, and criteria of included studies. The prevalence of hypertension among people diagnosed with diabetes was pooled in a mixed-effects meta-analysis using the “metafor” package in R. We used DerSimonian-Laird’s random-effects model as we anticipated considerable heterogeneity among the various studies. We applied the generalized linear mixed models along with logit transformation to determine the pooled prevalence across studies. (58). We also computed the 95% confidence interval (CI) for individual studies and the pooled prevalence using the Clopper-Pearson interval. The pooled prevalence of hypertension was stratified by five regions in Africa (East, West, North, South, and Central).

### Analysis

Pooled prevalence was calculated for hypertension in people diagnosed with diabetes. We used the random-effects model as we anticipated considerable heterogeneity among the various studies. We applied the generalized linear mixed models along with logit transformation to determine the pooled prevalence across studies (58). We also computed the 95% confidence interval (CI) for individual studies and the pooled prevalence using the Clopper-Pearson interval.

### Quality and Risk of bias Assessment

TH and AS independently conducted quality assessments using a Mixed-Method Appraisal Tool (MMAT)(16). We initially applied the first two mandatory questions to ascertain the feasibility of the tool. We used the standardised set of questions, Q4.1-Q4.5 which is designed for cross-sectional study designs. The overall quality score of the studies was the cumulative average of the independent scores of the two reviewers. Studies that met 4 or 5 of the quality assessment criteria were adjudged to be of high quality and low risk of bias. A rating of 3 means the study met three of the quality assessment criteria, and medium quality and low risk of bias. However, studies rated 2 or 1 showed poor quality and had a high risk of bias **(Table 2)**.

## Results

### Study results and identification

The initial electronic search yielded 3807 records. De-duplication led to exclusion of 564 articles, the 3243 articles progressed through titles and abstracts screening, with the exclusion of further 3126 records. The full texts of the remaining 117 records were extensively reviewed and guided by the eligibility criteria. Three additional articles were identified through hand searching of reference lists. The updated search yielded an additional five studies, published between 2021-2022. Overall, 41 articles fully met our inclusion criteria and were included in this review and the meta-analyses. Details of the screening process are provided in the PRISMA flowchart in **Figure 1**.

**Figure 1.**
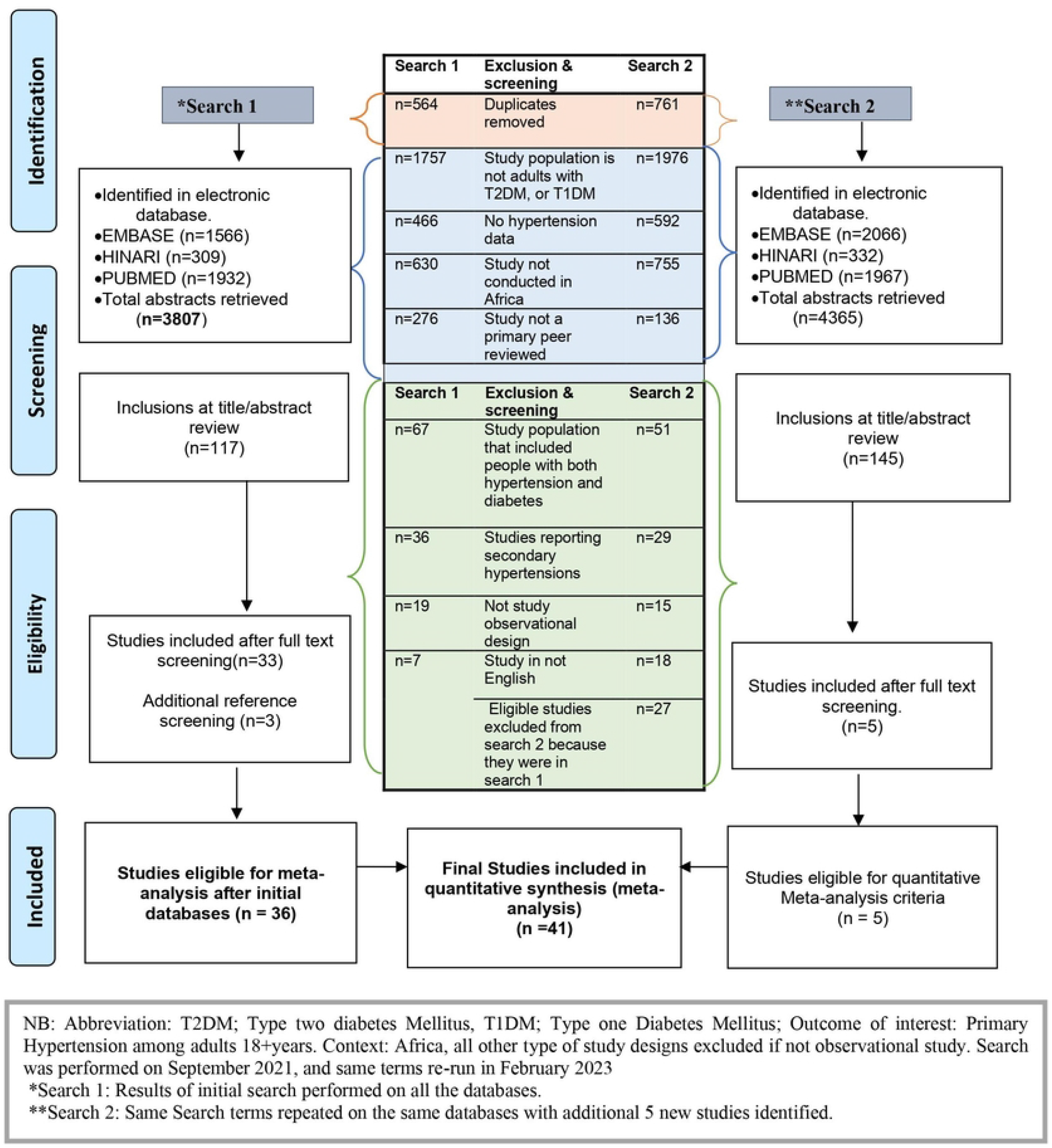

### Characteristics of included studies

Studies were representative of the five regions in Africa. There were more studies conducted in East, West, and South Africa than the North and Central Africa. Included studies were published between 2002 and 2022. **Figure 2** shows the burden of hypertension among people diagnosed with diabetes according to regions in Africa. The sample sizes of the studies ranged from 80 – 116726 and the mean age of the participant was 58 (± 11).

**Figure 2.**
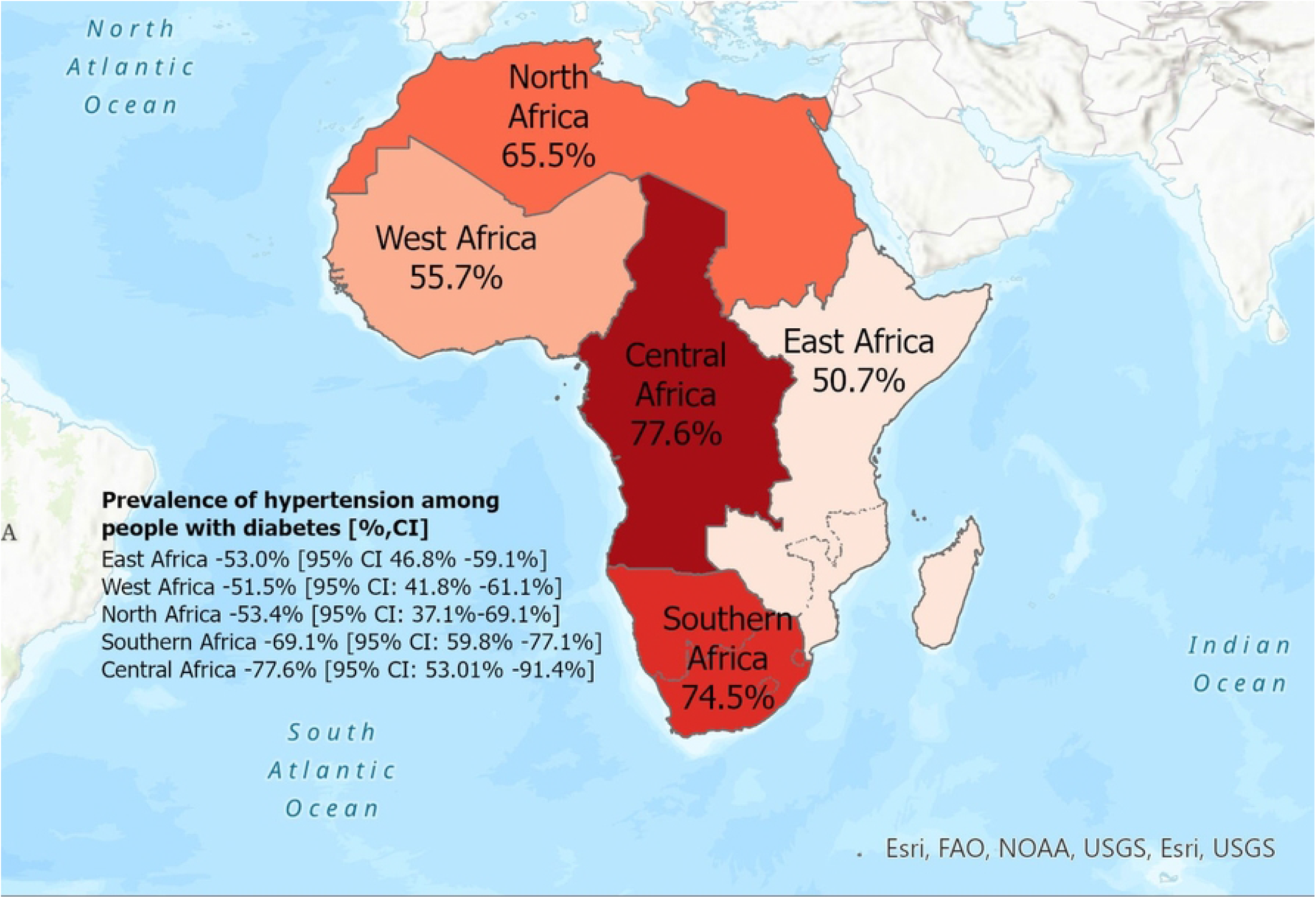

While most 32 (78%) of the studies utilized a cut-off point of ≥140/90 mm Hg for hypertension diagnosis, some 3 (8%) used ≥130/80 mm Hg as the cut-off. A few of the studies included in the review used an A1c threshold between (≥5.7 and >7.0) (17,19–21,24–27,29,30,32,34,36,42,43,45,49,52,55). In some cases, a fasting plasma/blood glucose level of 5-7 mmol/L or more was used (34,57). Other studies used HbA1c levels exceeding 6.5 or 7 as a diagnostic criterion for diabetes (17,19,29,30,39,42,49,52,54,55). While Twenty-four studies (58%) included patients diagnosed with only type 2 diabetes, no study was found among patients with only type 1 diabetes. Overall, the prevalence of hypertension among people with diabetes ranged from 21% in a study conducted in Ghana (34) to 92% in a South African sample (52).

### Prevalence of hypertension in patients with diabetes (Type 1 and 2)

Forty-one studies had complete data on the prevalence of hypertension among people diagnosed with diabetes and hence were eligible for inclusion in the meta-analysis. Overall, the prevalence of hypertension among people diagnosed with diabetes was 58.1% (95% CI: 52. % - 63.2%) shown in **Figure 3**. There was significant statistical heterogeneity across studies (i^2^ = 98%, p<0.001). The data were grouped into the five African sub-regions. The prevalence of hypertension among people diagnosed with diabetes within East, West, North, Central, and South Africa was 53.0% (95 CI: 45.8% - 59.1%; i^2^ = 96%, p<0.01), 51.5% (95 CI: 41.8% - 61.1%; i^2^ = 97%, p<0.01), 63.4% (95 CI: 37.1% - 69.1%; i^2^ = 99%, p<0.01), 77.6% (95 CI: 53.0% - 91.4%; i^2^ = 92%, p<0.01) and 69.1% (95 CI: 59.8% - 77.1%; i^2^ = 100%, p<0.01).

**Figure 3.**
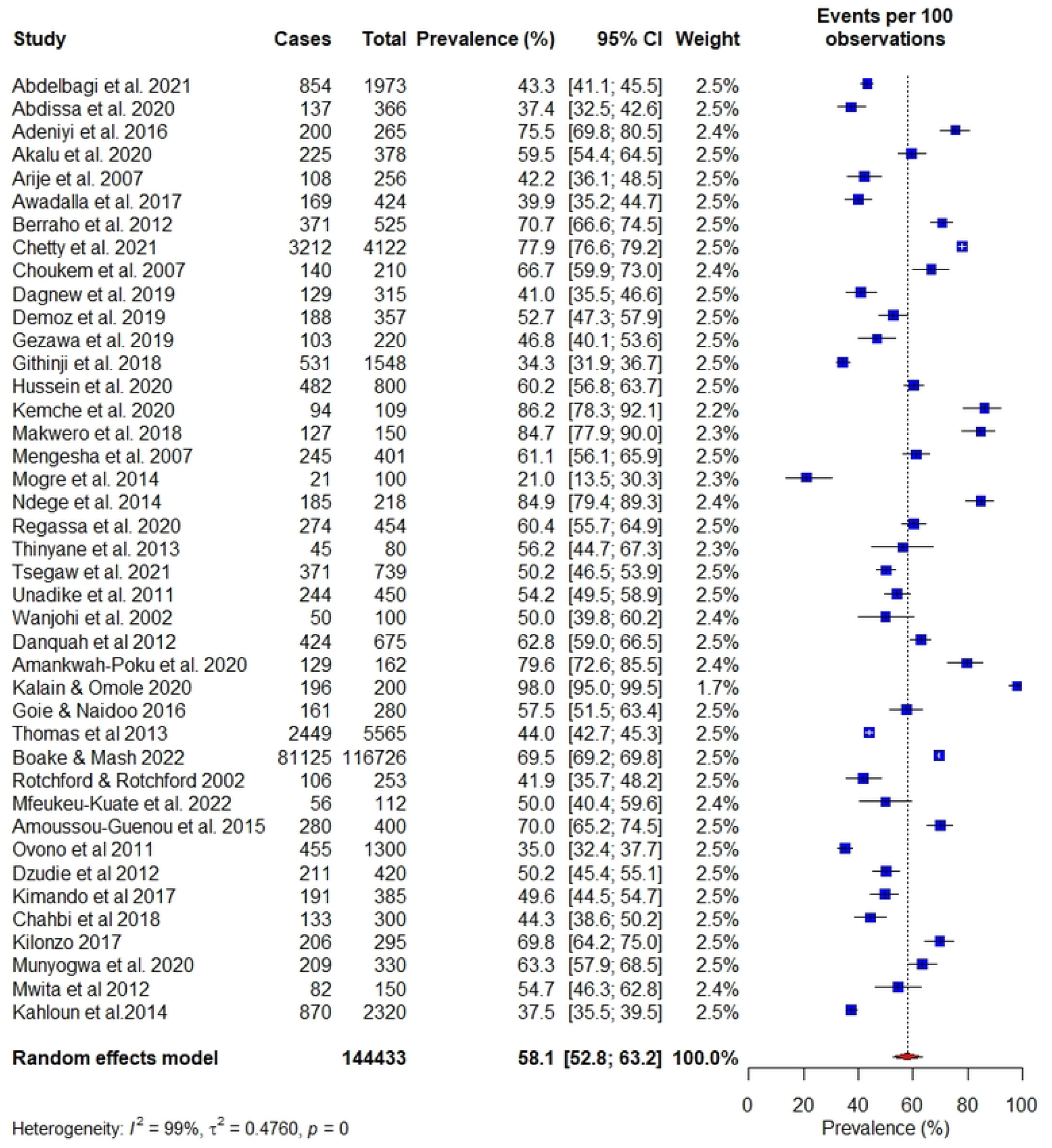

### Prevalence of hypertension in patients with only type 2 diabetes

Twenty-four studies (59%) included people diagnosed with type 2 diabetes **(Figure 4)**. Subgroup analysis among those with type 2 diabetes showed a prevalence of 60.8% (CI:53.4%-67.7%; i^2^ =99%, p<0.01) which was higher than the overall prevalence rate among T1DM and T2DM “combined” (58%). The highest prevalence was in South Africa (92%) (among patients with known hypertension at an outpatient department), followed by Ethiopia (84.9%), Zimbabwe (80%), and Ghana (74.6%) among participants with only T2DM. Some countries had only one study, hence estimation of the lowest prevalence among people with T2DM was impossible. However, we presented the prevalence the hypertension prevalence among patients with T2DM by the regions with countries within the African sub-region ranked according to the highest estimate **(Figure 5)**.

**Figure 4.**
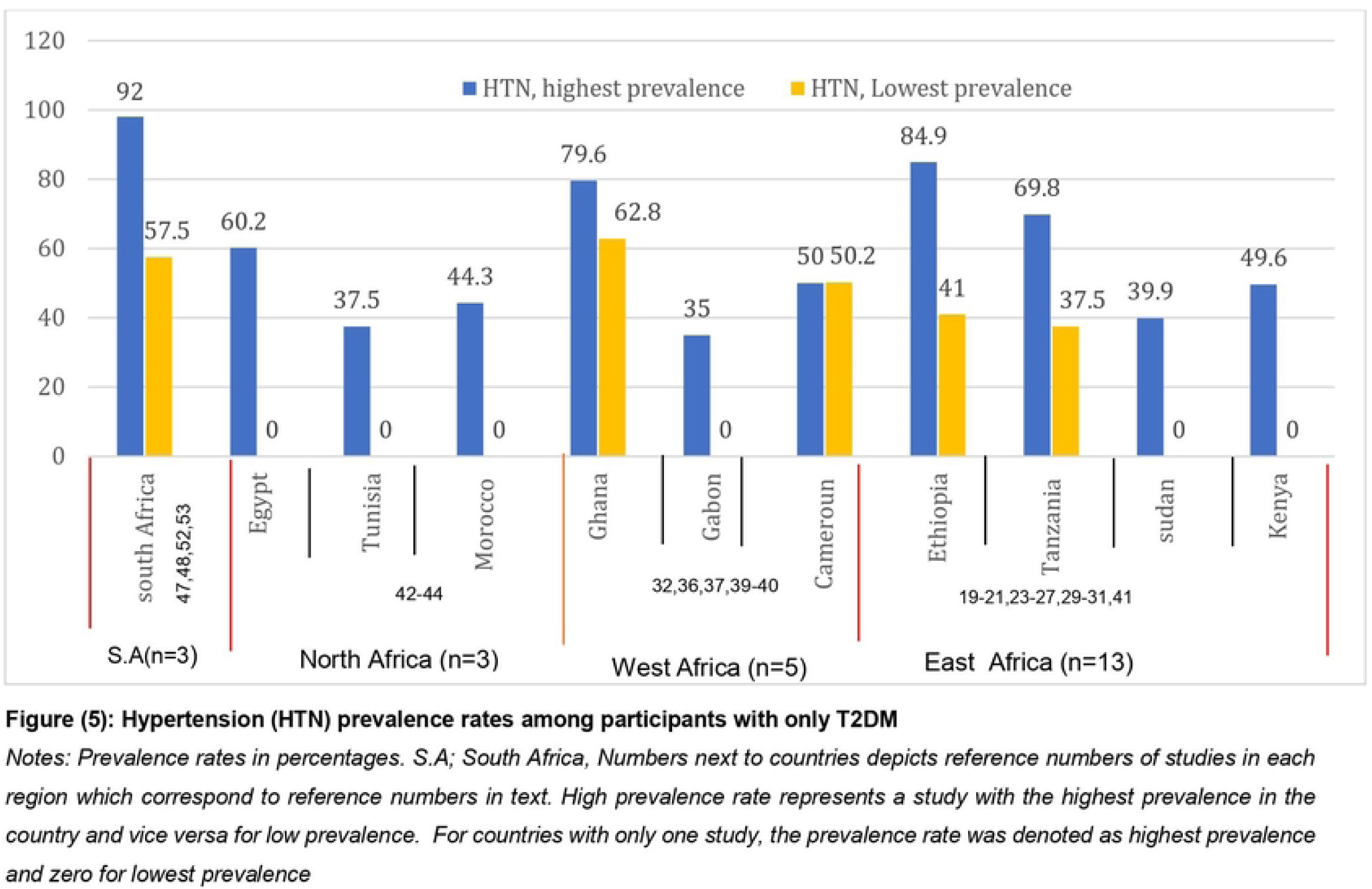
Prevalence rate accorting to country.

### Factors associated with hypertension among people with diabetes (Type 1 and 2)

Some studies reported factors associated with developing hypertension among people with diabetes. For instance, Abdellagi and colleagues found that the odds of developing hypertension increased with increasing age, body mass index, and being employed. In one study, age increased the odds of hypertension by more than 600% (OR 7.26 4.20-12.54) (41). Furthermore, the risk for hypertension among people with diabetes is highest within the 50-60 age category (29). The duration of diabetes is also associated with an increasing risk of developing hypertension. In three studies, being male increases the odds of developing hypertension among people diagnosed with diabetes. With regards to the residence, Akalu et al. (2020) and Awadalla et al. (2017) reported that people diagnosed with diabetes living in urban residence have higher chances of developing hypertension.

## Discussion

Our study reviewed the available literature and conducted a meta-analysis on the burden of hypertension among people with diabetes in Africa. It is well established that diabetes and hypertension are common conditions that are likely to coexist in people through multifaceted and multifactorial pathophysiology (59). In this study, more than half (58.1%) of the people with diabetes in African countries have hypertension. The burden is higher even among patients of known hypertension status and receiving treatment. For instance, 92% of patients diagnosed with diabetes accessing healthcare in an outpatient department in South Africa had hypertension. Significantly, few studies included in this review had higher hypertension prevalence among patients accessing care in health facilities. Furthermore, the prevalence of hypertension among people with only type 2 diabetes is significantly higher than type 1 and type 2 combined. This is suggestive of the rising burden of hypertension among people with type 2 diabetes in Africa. While this highlights the need for further research to explore the physiological mechanism between the risk of hypertension among people with type 2 diabetes, the few studies that included type 1 diabetes patients could be a result of a lack of capacity for screening and diagnosis of type 1 diabetes in Africa.

The findings of this review are particularly salient in light of micro and macrovascular complications and the state of the healthcare system in parts of Africa. In a systematic review by Colosia et al. which included seven studies from three African countries, the prevalence of hypertension among patients diagnosed with diabetes ranged from 38.5-80% which is consistent with the findings of our review (60). Additionally, factors such as age (50-60 years), long duration of diabetes, high BMI, male sex, and living in urban areas were noted in our analysis to contribute to the burden of hypertension-diabetes co-morbidity. A systematic review by Bosu and colleagues in 2021 reported an estimated prevalence of 57% of hypertension among the general adult population (>50 years) in Africa (61). Though their review did not exclusively include people diagnosed with diabetes, it highlights the increasing prevalence of hypertension with a higher BMI and above age 50 years, which frequently are the highest indicators of the development of diabetes or undiagnosed diabetes. These factors also corroborate with the findings of the largest United Kingdom Prospective Diabetes Study (UKPDS) in 1998 which reported an association between age, increased BMI, bigger waist/hip ratio, sedentary lifestyle with diabetes development, and further cardiovascular risk (62).

Overall, this review has significant implications for health systems’ orientation to caring for hypertension and diabetes in Africa. Firstly, this review highlights the need for an integrated NCD model of care in Africa. An integrated NCD model of care is a cost-effective opportunity to prioritize the healthcare needs of people diagnosed with both hypertension and diabetes in settings where healthcare workers and resources are limited.

Secondly, the higher prevalence of hypertension reported in this review can be attributed to concerns about urbanization which impact the behavioural styles of the populace. Community and clinic-based models of screening are potential opportunities to be leveraged to enhance screening, treatment, and early detection of hypertension and diabetes.

### Strengths and limitations

Previous reviews have focused on hypertension or diabetes burden in infectious diseases such as HIV/AIDS and Tuberculosis in Africa (64–66). However, this study is the first review to estimate the burden of hypertension among people diagnosed with diabetes in Africa. Additionally, this review captured the prevalence rate among people with type 2 diabetes as well as type 1 diabetes. However, the review needs to be considered in the context of some limitations. Only studies published in English were included and that may cause language bias, especially as some African countries publish in other languages like French. More specifically, countries in central Africa were likely to be under-represented which could affect the prevalence estimate. Nonetheless, this review provides a snapshot of the rising burden of hypertension among people with diabetes in Africa.

## Conclusion

Hypertension and diabetes co-morbidity is high in Africa. Affected individuals are at high risk for both microvascular and macrovascular complications of diabetes, which include retinopathy, nephropathy, and neuropathy (microvascular) whereas the macrovascular ones include the diseases of the coronary arteries, peripheral arteries, and cerebrovasculature. Patients with these co-morbidities should be managed with lifestyle modifications and appropriate medications to reduce the risk of developing these associated complications.

## Data Availability

All data used in this manuscript has been included in this manuscript

